# Transformers for rapid detection of airway stenosis and stridor

**DOI:** 10.1101/2024.10.17.24315634

**Authors:** James Anibal, Rebecca Doctor, Micah Boyer, Karlee Newberry, Iris De Santiago, Shaheen Awan, Yassmeen Abdel-Aty, Gregory Dion, Veronica Daoud, Hannah Huth, Stephanie Watts, Bradford J. Wood, David Clifton, Alexander Gelbard, Maria Powell, Jamie Toghranegar, Yael Bensoussan, the Bridge2AI Voice Consortium

## Abstract

Upper airway stenosis is a potentially life-threatening condition involving the narrowing of the airway. In more severe cases, airway stenosis may be accompanied by stridor, a type of disordered breathing caused by turbulent airflow. Patients with airway stenosis have a higher risk of airway failure and additional precautions must be taken before medical interventions like intubation. However, stenosis and stridor are often misdiagnosed as other respiratory conditions like asthma/wheezing, worsening outcomes. This report presents a unified dataset containing recorded breathing tasks from patients with stridor and airway stenosis. Customized transformer-based models were also trained to perform stenosis and stridor detection tasks using low-cost data from multiple acoustic prompts recorded on common devices. These methods achieved AUC scores of 0.875 for stenosis detection and 0.864 for stridor detection, demonstrating potential to add value as screening tools in real-world clinical workflows, particularly in high-volume settings like emergency departments.

## 1. Introduction

Upper airway stenosis is defined as a narrowing of the airway which can occur at various levels within the laryngo-tracheal apparatus and can be caused by factors ranging from abnormal scar tissue, traumatic injuries, malignancy, or neurological paralysis of the vocal folds. When the narrowing is significant, airway stenosis often results in stridor, a respiratory bio-acoustic signal caused by turbulent airflow and tissue vibration at different levels within the upper airway.^1-2^ Vibrating tissue and turbulent airflow through the human airway create unique acoustic signals.

In previous literature, stridor has been characterized as inspiratory, expiratory, or biphasic, with some proposed correlation to the level of the obstruction.^4^ Stridor has also been classified by anatomical location, including supraglottic, glottic, subglottic, extra-thoracic, and intrathoracic.^5^ Recognizing stridor is an important clinical skill, as trained experts will listen to stridor and patient respiratory effort to assess the severity of different airway pathologies and determine the appropriate intervention. For example, presence of inspiratory stridor is used to differentiate severity and treatment for patients with parainfluenza virus causing subglottic swelling.^6^ Acute onset stridor may indicate critical decompensation of a patient’s upper airway patency, which can be life-threatening if not urgently identified and treated, such as in cases of epiglottitis or anaphylaxis.^7^ Stridor can also act as a clinical indicator for intubation, or in assessment of post-extubation laryngeal edema. ^8^

The misdiagnosis of airway stenosis can delay onset of critical treatment interventions. An informal poll conducted on an international Facebook group for over 8,200 patients with airway stenosis found that 96% of the participants reported being initially misdiagnosed, causing extended periods of unresolved health challenges/risk.^9^ Stridor is a common symptom of airway stenosis, but is often mistaken as wheezing in clinical practice, resulting in frequent misdiagnosis as asthma.^10^ Failure to recognize stridor as a sign of severe airway stenosis can lead to important morbidity and mortality. Patients with stridor may need emergent interventions or, if undergoing procedures, customized intubation with technologies like specialized fiberoptic laryngoscopes.

The gold standard for investigating pathologies causing airway stenosis and potentially stridor is through direct visualization by laryngoscopy and bronchoscopy.^11^ Unfortunately, many misdiagnosed patients are not referred to expert otolaryngologists promptly, which can result in significant, high-risk delays in the onset of care. Moreover, many otolaryngologists do not perform in-office laryngoscopies below the level of the glottis and may miss subglottic and tracheal stenosis.

Recent advances in machine learning offer the means to potentially recognize unique acoustic signals from different pathologies. Machine learning models may be more sensitive than the human ear and might be trained to identify airway stenosis and stridor, enabling non-invasive screening methods or decision-support tools for healthcare professionals. However, due to challenges in data acquisition and annotation, limited work has been completed in this space compared to areas such as cough assessment with AI models.^12^ This work makes the following contributions:

### A unified dataset to train AI models for detection of airway stenosis and stridor

The dataset used in this study contained data from patients with upper airway stenosis, stridor, other respiratory/voice conditions, and general controls. The records in this dataset were collected non-invasively from low-cost point-of-care devices via tasks which can be rapidly completed and are not dependent on literacy.

### Airway stenosis detection with deep learning

a customized transformer model was trained to perform stenosis detection using YAMNet representations of forced inhales and normal deep breaths, offering a potential solution to the challenging problem of misdiagnosis.

### Stridor detection with deep learning

a second transformer model was trained to identify stridor from YAMNet embedding matrices. This model may be used with the airway stenosis model in a hierarchical system for additional severity assessments (e.g., pre-intubation) or independently for the detection of stridor in cases which may not involve upper airway stenosis but may still be life-threatening.

## 2. Related Work

There have been relatively few successful attempts to classify airway stenosis with machine learning. Past experiments were mainly centered around small datasets collected from invasive or high-cost modalities such as electromyography, spirometry, and imaging studies.^13-16^ Similarly, there has been limited work on the use of AI to detect stridor. One recent effort utilized video-polysomnography data and few-shot learning to detect stridor in a small number of patients (n=18).^17^ This approach achieved over 96% detection accuracy using data from only eight stridor patients for training the model, outperforming a state-of-the-art AI baseline by 4%–13%.^17^ A different study attempted to use power spectrums and neural networks to classify 9 different categories of respiratory sounds, one of which included stridor.^18^ This was done on a dataset of 36 patients collected via online sources and a high-cost electronic stethoscope, with only two patients per class in the training set.^18^ Although progress has been made in advancing stridor detection with AI, further advancements are necessary to classify stridor using more scalable data collection techniques like acoustic recordings from smartphones or other low-cost devices.

## 3. Methods

In this section, datasets, preprocessing methods, machine learning techniques, and validation strategies are described in detail. Table 1 provides definitions for key clinical terminology used throughout this report.

**Table 1.**
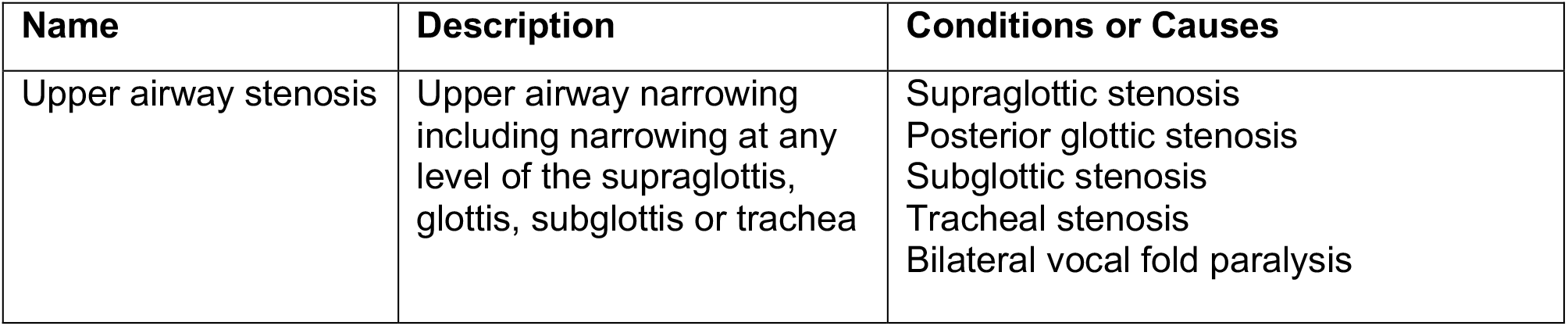
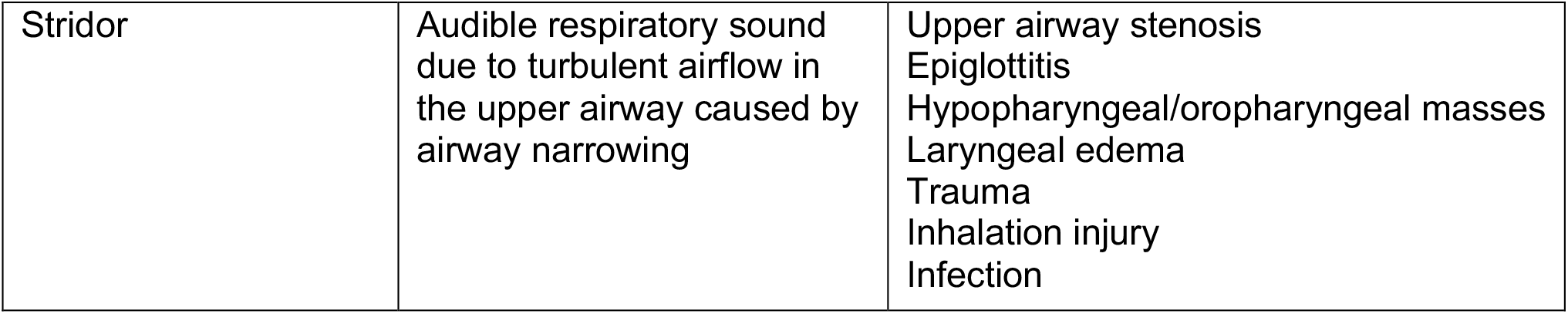
Definition of clinical terms referenced in this study.

### 3.1 Datasets

The data used in this study comes from two main sources: a dedicated stenosis/stridor dataset (“USF Stridor”) collected from the USF Health Voice Center at the University of South Florida (USF) and the Bridge2AI Voice Dataset from the Bridge2AI Voice Data Generation Project (“Bridge2AI”).^19^ In this study, the Bridge2AI dataset was used for additional airway stenosis records and the diversification of the control cohort. The unified dataset contained 94 patients in the airway stenosis cohort, 45 patients in the stridor cohort, 51 patients in the cohort of other disordered voice/respiratory disease controls, and 77 patients in the general control cohort (no medical conditions or unknown health history).

The disordered voice and respiratory disease control cohort contains patients with the following conditions: recurrent respiratory papillomatosis (RRP), COPD, Asthma, Spasmodic Dysphonia, Obstructive Sleep Apnea (OSA), Chronic Cough, Benign Vocal Cord Lesion, Laryngeal Cancer, and Vocal Fold Paralysis. Table 2 contains demographic information about each cohort in the dataset (there is an overlap between the stenosis and stridor cohorts).

**Table 2.** Demographic Statistics from the unified dataset (USF Stridor and Bridge2AI)

| Cohort | Male | Female | Other N/A | 18-34 | 34-50 | 50-64 | 65+ | N/A |
| --- | --- | --- | --- | --- | --- | --- | --- | --- |
| Airway Stenosis (Pathology) | 24 | 69 | 1 | 14 | 8 | 41 | 29 | 2 |
| Stridor (Sound) | 9 | 36 | 0 | 7 | 2 | 20 | 15 | 1 |
| Other Respiratory/Voice | 20 | 30 | 1 | 4 | 6 | 15 | 24 | 2 |
| General Controls | 30 | 41 | 6 | 28 | 11 | 10 | 28 | 0 |

#### 3.1.1 Acoustic Tasks

In this study, AI models were trained using audio data collected from two different acoustic tasks (Table 3): forced inhale with the mouth open (FIMO) and normal deep breaths (DB). The tasks were chosen by expert practitioners and laryngologists for multiple reasons, including simplicity, no dependency on literacy or English fluency, elimination of nasal interference due to nasal turbulent airflow (often caused by seasonal allergies or other benign factors), and increased sensitivity to mild stridor due to the forcible pushing of air over the vocal cords (FIMO), increasing the likelihood of audible disordered sound. Before completing these tasks, participants were given the following instructions:

**Table 3.**
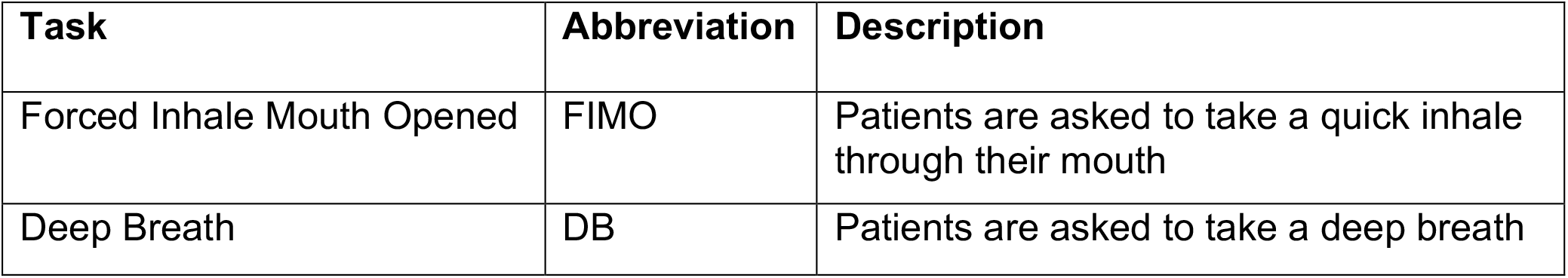
Acoustic tasks used to collect stridor sounds.

| Task | Abbreviation | Description |
| --- | --- | --- |
| Forced Inhale Mouth Opened | FIMO | Patients are asked to take a quick inhale through their mouth |
| Deep Breath | DB | Patients are asked to take a deep breath |

##### Deep Breath Data

Take 5 big breaths in and out through your mouth.

##### Forced inhales through the mouth

Exhale, then inhale quickly through your mouth, as if you are trying to catch your breath. Record three of these breaths in a single recording.

#### 3.1.2 Data Collection Devices

FIMO and DB recordings were collected from various technologies depending on the study (USF stridor or Bridge2AI) and the availability of devices. These included an At2O35 AudioTechnica microphone placed at 12 inches from the mouth, a previously validated low-cost Avid AE-36 headset microphone placed at 2-3 inches from the mouth, and an internal iPad microphone placed at 6 inches from the mouth.^20-21^ To achieve generalizable AI methods which can be deployed in real-world settings, compatibility with multiple devices is essential due to the differences in technological ecosystems across different healthcare centers.

For patients in the USF Stridor study, audio data was collected from the full range of recording technologies described above (if possible). However, due to limitations related to the availability of devices and patient compliance, this was not always feasible, demonstrating the need for scalable solutions which can accommodate different types of data. If data from multiple devices was available for one patient, all the data was retained to ensure the maximum number of audio samples were available for model training. These different devices produced similar (but not identical) data points, which may be comparable to audio data augmentation techniques like spectrogram masking or the addition of random noise.^22^ For the controls in the Bridge2AI Voice dataset, audio data was collected using only the Avid headset microphone placed 2-3 inches from the mouth. Table 4 includes the quantities of data collected from different microphones.

**Table 4.** Statistics on microphone types and acoustic tasks within the cohorts included in the training datasets (there was overlap between the stenosis and stridor cohorts).

| Cohort | Device | FIMO | DB |
| --- | --- | --- | --- |
|  | At2O35 | 314 | 0 |
| **Airway Stenosis (Pathology)** | Avid Headset | 1970 | 4590 |
|  | iPad | 575 | 755 |
|  | At2O35 | 208 | 0 |
| **Stridor (Sound)** | Avid Headset | 1134 | 1768 |
|  | iPad | 499 | 594 |
|  | At2O35 | 0 | 0 |
| **Respiratory Controls** | Avid Headset | 1141 | 2989 |
|  | iPad | 0 | 0 |
|  | At2O35 | 94 | 0 |
| **General Controls** | Avid Headset | 1596 | 4608 |
|  | iPad | 299 | 379 |

#### 3.1.3 Data Annotations

Clinical metadata (patient and physician reporting) was then used to determine if the patients had upper airway stenosis or any other respiratory conditions (the latter being a key consideration in the curation of robust control cohorts). The presence or absence of stridor was determined by a laryngologist with expertise in airway stenosis. These annotations were based on respiratory phase (inspiration/expiration) and the presence of audible stridor (stridor or no stridor). Due to the use of a single annotator, which may more directly resemble scalable clinical data collection processes for real-world environments (particularly in resource-constrained settings), this data was considered weakly supervised.^23^ The FIMO data was used as the basis for annotation due to the higher likelihood of stridor detection during this type of breathing compared to other tasks. For experiments involving stridor detection, patients with airway stenosis but no audible stridor were included in the cohort of controls with other respiratory conditions.

Deep breathing recordings collected during the same recording session as the FIMO task were assumed to have the same stridor status as the FIMO data. This resulted in weakly supervised “inferred annotations” which may present an additional source of regularization for training deep learning models, possibly enhancing downstream performance.^23^ However, only the FIMO data was used for validation and testing of the stridor detection models. Other tasks were not directly assessed for the presence of stridor, and the inferred annotations were not considered sufficiently robust to be included in the model performance metrics.

#### 3.1.4 Data Quality Control

Acoustic data was excluded from the study if the patient was fully non-compliant with the instructions (e.g., phonated an elongated vowel instead of deep breathing). For the stenosis and stridor cohorts, data was also removed if any errors were made in the data collection protocol (e.g., breathing through the nose) that could have obscured true biomarkers of disease. The control cohorts were not subjected to this second phase of filtering, potentially introducing noise into the dataset which may be similar to disordered breathing. Cohort-specific quality control pipelines were applied to increase the likelihood that more challenging negative cases would be shown to the model during training. For example, a healthy patient may phonate a gasping sound when asked to forcibly inhale. This can result in a stridor-like sound unrelated to airway obstruction. The presence of such data in the training set may enhance downstream generalization by encouraging the model to learn nuanced, disease-specific representations which enable separation between actual pathological signals and benign variations.

### 3.2 Data Preprocessing

The FIMO and DB audio signals were broken into 2.5-second segments of raw audio waveforms, with 80% overlap. The segment length and overlap percentage were based on the timing of breathing cycles to ensure that some component of the inhale signal was present in all the data points: exhales are much less likely to contain stridor or other biomarkers of airway narrowing during FIMO and DB acoustic exercises. The 80% overlap was further used as a form of data augmentation, to expand the number of “different” audio segments available for training AI models. The preprocessing steps described here resulted in a training dataset of 19310 audio segments in total. The number of segments per microphone, task, and cohort can be found in Table 4.

### 3.3 YAMNet Embeddings

After initial preprocessing, the 2.5-second audio segments were standardized to a sampling rate of 16.1 kHZ, normalized to the range of [-1,1], converted into Mel spectrograms, and encoded by the YAMNet model.^24-25^ YAMNet was chosen due to extensive pre-training on millions of YouTube audio segments from 521 categories of sound.^23^ This domain knowledge ensures that the model can encode nuanced differences between sounds and may enhance performance on tasks involving breathing audio data with subtle acoustic biomarkers of disease. The YAMNet model returns a matrix containing one embedding vector for each 0.5 second window of data (5*x*1024 dimensions for a 2.5 second segment). For this study, YAMNet was also used as part of the data filtering process. After generating the embedding matrix, the predicted probabilities for the sound type were output by the classification head of the model. If more than 50% of the segment was placed in a category which indicated a potentially significant source of external noise or microphone malfunction, such as electronic buzzing or humming, the segment was removed from the dataset.

### 3.4 Model Training

A transformer model was trained on the YAMNet embedding matrices to detect airway stenosis or stridor (Fig. 1).^26^ Transformers rely on global self-attention mechanisms to capture the key relationships (between embedding vectors in the matrix) to which the model should “pay attention” and have demonstrated robust performance on advanced tasks with similar data structures (e.g., matrices of word or image embeddings).^26-28^ Unlike conventional transformers for natural language processing or computer vision, positional encodings were not used within the model architecture. This minor adaption of the original transformer algorithm was done to ensure translational equivariance for audio data points with similar features at different positions in the matrix of YAMNet embedding vectors. Due to the application of data chunking with overlap, absolute position would not be relevant and may cause model confusion.

**Figure 1.**
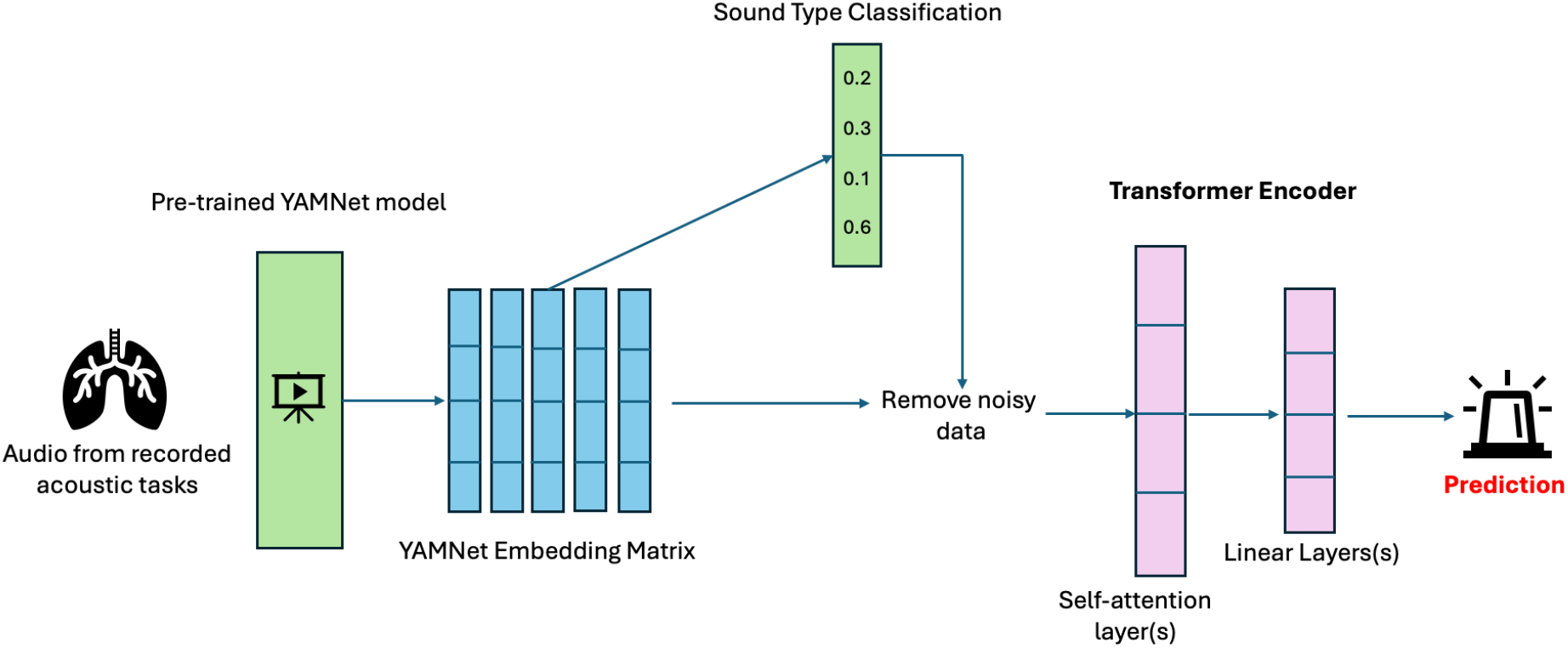
The data processing and modeling pipeline for stenosis and stridor detection.

An adaptive movement estimation function was used to train the model based on the cross-entropy loss calculated from minibatches of data. Early stopping was used to optimize training based on the validation loss. No distinctions were made between data from the different recording devices included in the experiments (Avid, AtO235, iPad).

The workflow shown in Figure 1 includes (1) the generation of YAMNet embedding matrices from Mel Spectrograms, (2) the filtering of noisy data based on sound type classification, and (3) the use of a transformer encoder to predict the label of the input data.

### 3.5 Validation

Nested *k*-fold cross validation was used to estimate the performance of the transformer models, ensuring a credible evaluation of generalization potential. The prediction threshold for the test data was determined by aggregating results from the *k* validation sets. If a test patient had data available from multiple recording systems, the prediction was made based on the best available device in terms of representation within the training dataset (by number of samples). For this study, data from either the Avid headset, the iPad, or the At2O35 AudioTechnica, respectively, were considered in the final prediction. Here, the aim was to simulate the real-world case wherein only a single device (presumably the best device available at that time) would be used to collect data from a patient – multiple devices would not be used due to time/resource limitations. For stenosis detection, if a test patient had available data from multiple acoustic exercises (FIMO, DB), inference was run separately, and the mean probabilities were calculated for each task. The average of these probabilities was used for decision-making purposes. In the experiments involving stridor detection, only FIMO data was used in the validation/test sets due to the nature of the manual annotation (DB was not checked for stridor).

## 4. Results

In this section, results are presented for transformer models which were trained to complete two tasks from recorded breathing data: (1) the detection of airway stenosis and (2) the detection of stridor. Potential use cases for these models are described in Table 5. All performance metrics are reported based on the mean result across multiple iterations of the experiment, accounting for variability due to the random initialization of transformer weights.

**Table 5.**
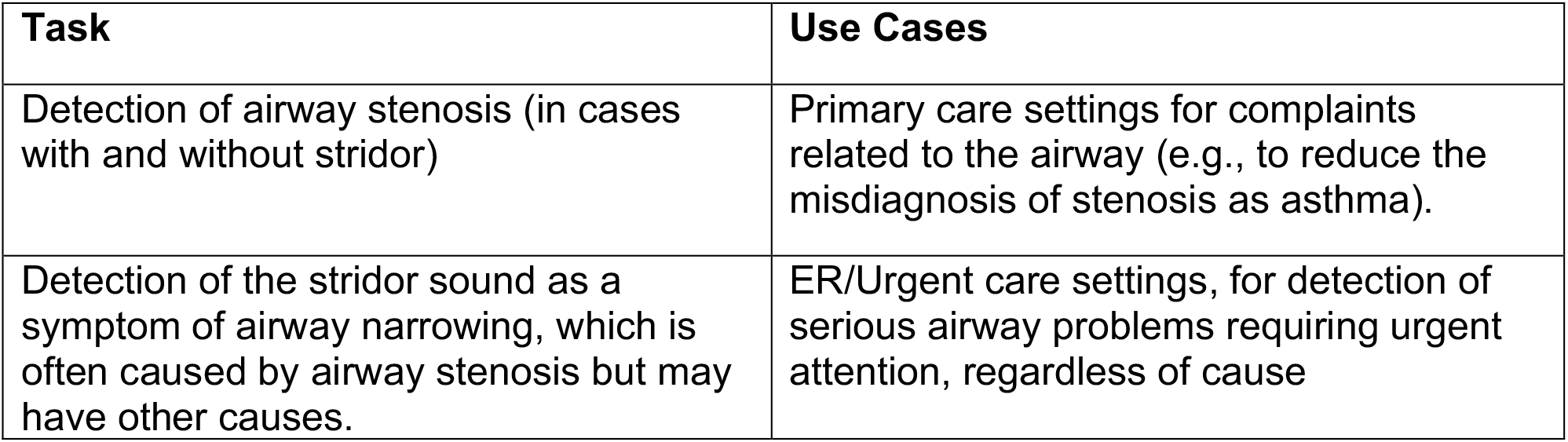
Potential use cases for AI models trained within this study.

| Task | Use Cases |
| --- | --- |
| Detection of airway stenosis (in cases with and without stridor) | Primary care settings for complaints related to the airway (e.g., to reduce the misdiagnosis of stenosis as asthma). |
| Detection of the stridor sound as a symptom of airway narrowing, which is often caused by airway stenosis but may have other causes. | ER/Urgent care settings, for detection of serious airway problems requiring urgent attention, regardless of cause |

Table 6 shows the sensitivity (calibrated to a value of 0.85 in consideration of the life-threatening nature of airway failure), specificity (at 0.85 sensitivity) and the AUC score. The life-threatening nature of airway obstruction justifies the prioritization of sensitivity in this case. Results are presented using data from the “best available” recording device for a test patient (as described in section 3.5), mirroring the likely scenario where multiple sites have deployed the same AI model, but different devices are used depending on technology resources. If, for a patient in the test set, audio from the Avid headset was available, results are reported based on predictions made from this data. Otherwise, results are reported from the iPad or At2O35 recordings (in that order), favoring optimal data while accounting for potential technology limitations in real-world clinical environments. For specificity, three values are reported: overall, general control cohort, and respiratory control cohort (as defined in Section 3.1).

**Table 6.** Results from Airway Stenosis Detection and Stridor Detection Tasks.

| Task | AUC | Sensitivity | Specificity @0.85 sensitivity |
| --- | --- | --- | --- |
| Stenosis Detection | 0.875 | 0.851 | 0.725 (0.792/0.624) |
| Stridor Detection | 0.864 | 0.856 | 0.684 (0.742/0.624) |
| Stridor Detection<br>(FIMO only, no DB data) | 0.846 | 0.852 | 0.661 (0.703/0.617) |

### 4.1 Detection of Airway Stenosis

The experimental results of this study show that the transformer model trained on YAMNet embeddings may have learned a moderately robust signal for the detection of airway stenosis (AUC of 0.875), with an overall specificity of 0.725 when sensitivity is calibrated to 0.85. Compared to other types of errors, the model is more likely to incorrectly classify other respiratory controls as having airway stenosis. Given the life-threatening implications of intubating a patient with airway stenosis and the 96+% misdiagnosis rate, this model may still represent a potentially deployable technology with real-world impact. ^9^

Figure 2 shows the difference in AUC score between different demographic groups within the dataset, addressing possible sources of bias in the training data. While the model was reasonably consistent, there were noticeable performance differences between age groups, possibly due to the differing age distributions between the control cohorts (Table 2). The general controls may be less challenging for the model compared to those with other respiratory conditions. The model also obtained a higher AUC score for female patients, which aligns with the distribution of upper airway stenosis cases in the dataset (Table 2) and in the real-world (the disease most often affects middle-aged women).^29^

**Figure 2.**
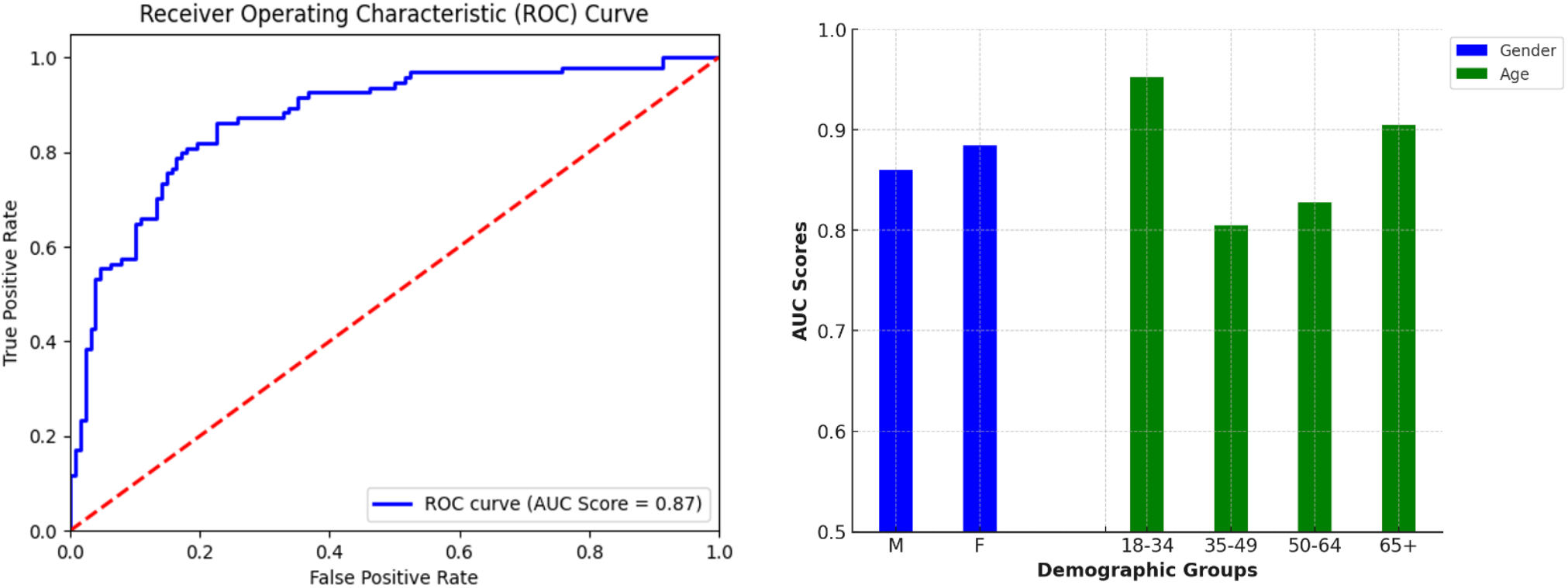
<u>Left:</u> mean ROC curve for the airway stenosis detection model. <u>Right:</u> Model performance (mean AUC score) across age and gender identity groups.

### 4.2 Detection of Stridor as a General Symptom of Airway Failure

A transformer model trained on YAMNet embedding matrices showed promising results on the stridor detection task as well, achieving an AUC score of 0.864 with an overall specificity of 0.684 (at 0.85 sensitivity). Expectedly, the performance of the model was weakest on the cohort of controls with other respiratory conditions, which included upper airway stenosis without audible stridor. Figure 3 (right) shows the difference in AUC score between different demographic groups in the dataset.

**Figure 3.**
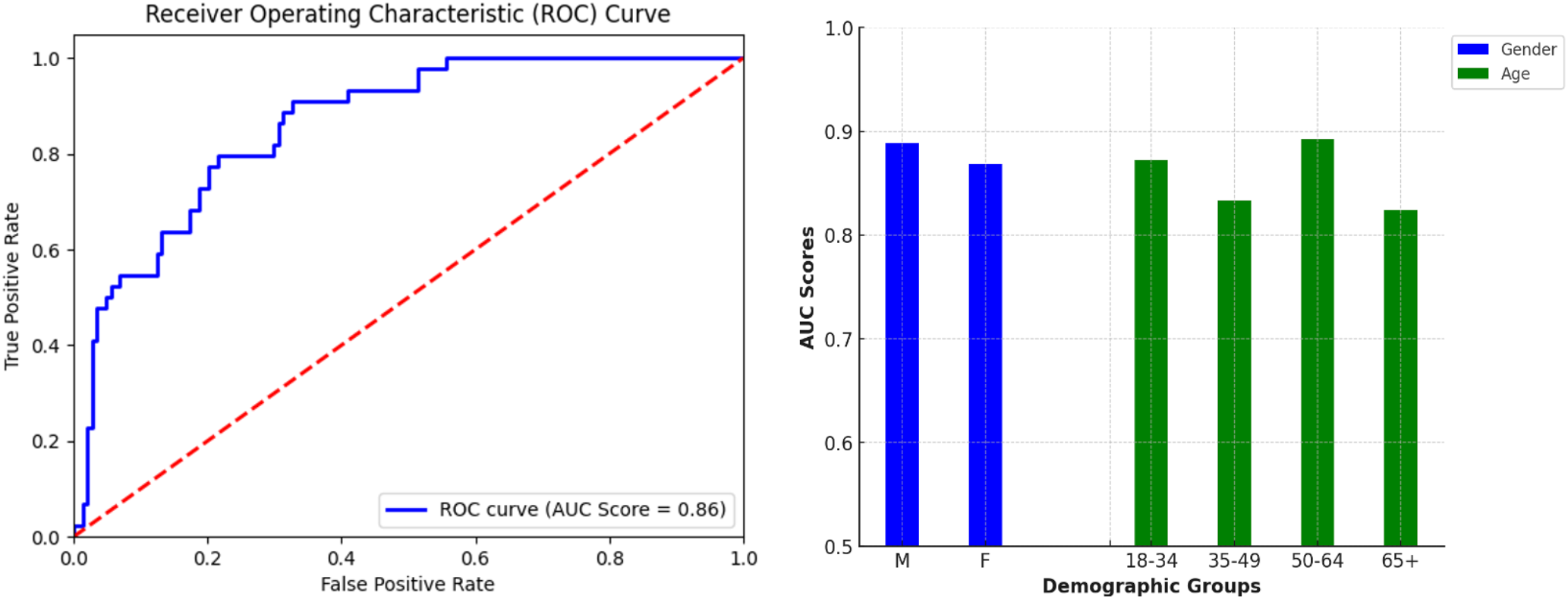
<u>Left:</u> mean ROC-AUC curve for the stridor detection model. <u>Right:</u> Model performance (mean AUC score) across age and gender identity groups.

Moreover, while only FIMO data was used in the validation/test sets, the transformer model which was trained on both FIMO and DB data achieved the best results. This was despite the weak “inferred” DB annotations in the training data. The transformer model trained without the DB data resulted in a lower AUC score of 0.846, with nearly a 4% difference in specificity for the general control cohort.

## 5. Discussion

In this report, a customized transformer model was trained to screen for airway stenosis and stridor using non-invasive data from multiple devices/acoustic tasks. A unified dataset was introduced for training the AI models, containing FIMO and deep breath records from two sources – the USF Stridor dataset and Bridge2AI Voice dataset. Upon completion of training, resultant AI models showed that there may be viable signal within low-cost, weakly annotated respiratory recordings, thereby facilitating the identification of airway stenosis and stridor cases. These findings may have relevance in the development of new digital health technology leveraging voice as a biomarker of health, with potential use cases in the ER, primary care/point of care settings, and voice health/laryngology clinics (Table 5).

### 5.1 Limitations

Promising results were obtained by this study, but there were multiple limitations which should be noted in the interpretation of the work. First, while larger than previous studies involving stridor and airway stenosis, the dataset was still limited – the stridor cohort contained 45 patients labeled by a single expert annotator, which may have implications for the generalizability of the weakly supervised AI system. The stridor annotations, while performed by an expert clinician, were done based on audio recordings alone, rather than multimodal data involving laryngoscope videos and EHR, which may have resulted in some instances of mislabeled data. The annotations were also binary, without information on the severity of the airway stenosis or stridor cases. Limited variations in audio recording equipment were used for data acquisition, and evolution of audio hardware may lead to distribution shifts affecting model performance.

In terms of performance, the models showed differences in AUC score between age groups and a slight skew towards female gender identity (though the conditions themselves more often impact female patients). Other potential biases like race/ethnicity, smoking status, and type of stridor (i.e., cases not caused by airway stenosis) must still be considered in future work. Moreover, the two datasets used in this study were collected in slightly different settings: USF Stridor data was recorded in a sound isolation booth, whereas the Bridge2AI data was recorded in a small, quiet room. This is not likely to cause a significant difference in results but may have introduced some false signal into the model due to the imbalance between the datasets (many of the controls came from the B2AI dataset). Finally, while the data and devices were scalable, the environment of the study may not directly replicate real-world conditions like an emergency room, which is likely to introduce additional sources of noise and more acute presentations of the disorders compared to the dataset used in this study.

### 5.2 Future Work

Future work will involve the design of a mobile application for deploying trained AI models on a smartphone/tablet, allowing healthcare professionals to test the viability of these tools in real-world settings. Robust performance in retrospective experimentation does not equate to utility in a clinical environment: implementation studies are essential to the improvement of the methods. Moreover, such an application may facilitate the collection of additional data across multiple types of airway disorders and different healthcare settings, including the emergency room.

Future work is also needed to correlate acoustic features with severity of the disease, including the percentage of stenosis in terms of airway circumference, the vertical length of stenosis, and severity of dyspnea. To achieve these results, acoustic signals must be correlated with measurements on CT scan imaging and video endoscopies/bronchoscopies or measurements collected during procedures (in the OR). A hierarchical implementation of the described AI methods may also add future value as a simultaneous predictor of both the presence and severity of airway stenosis (with stridor as indicator of severity). The work described here may also have potential for testing and deployment in low- and middle-income countries (LMICs), where airway failure is a significant challenge to healthcare providers in terms of technology and expertise.^30-31^ Rapid and low-cost AI-assisted support for airway stenosis or stridor detection in this context represents a novel opportunity.

## 6. Conclusion

Airway stenosis, and the stridor which often accompanies moderate to severe cases (or other life-threatening airway problems) is a challenging clinical issue related to misdiagnosis in healthcare. Misdiagnoses are common, which may worsen patient outcomes. Deep learning, which has proven capabilities in capturing nonlinear biomarkers of health, may be a solution for early identification and screening by general healthcare providers. In this study, transformer models were used to demonstrate the potential of clinically viable signal in the task of rapidly screening for airway stenosis and stridor using low-cost data. Future development of this work is essential to the downstream deployment of affordable, non-invasive tools for airway assessments in diverse, high-volume settings.

## **Members of the Bridge2AI Consortium who contributed to this work

Yael Bensoussan, University of South Florida, Tampa, FL, US; Olivier Elemento, Weill Cornell Medicine, New York, NY, USA; Anais Rameau, Weill Cornell Medicine, New York, NY, USA; Alexandros Sigaras, Weill Cornell Medicine, New York, NY, USA; Satrajit Ghosh, Massachusetts Institute of Technology, Boston, MA, USA; Maria Powell, Vanderbilt University Medical Center, Nashville, TN, USA; Vardit Ravitsky, University of Montreal, Montreal, Quebec, Canada; Jean Christophe Belisle-Pipon, Simon Fraser University, Burnaby, BC, Canada; David Dorr, Oregon Health & Science University, Portland, OR, USA; Phillip Payne, Washington University in St. Louis, St. Louis, MO, USA; Alistair Johnson, University of Toronto, Toronto, Ontario, Canada; Ruth Bahr, University of South Florida, Tampa, FL, USA; Donald Bolser, University of Florida, Gainesville, FL, USA; Frank Rudzicz, Dalhousie University, Toronto, ON, Canada; Jordan Lerner Ellis, University of Toronto, Toronto, ON, Canada; Shaheen Awan, University of Central Florida, Orlando, FL, USA; Stephanie Watts, University of South Florida, Tampa, FL, US; Jennifer Sui, Hospital for Sick Children, Toronto, Ontario, Canada; Karim Hanna, University of South Florida, US; Theresa Zesiewicz, University of South Florida, US; Robin Zhao, Weill Cornell Medicine, New York, NY, USA; Lochana Jayachandran, Mount Sinai Hospital, Toronto, Ontario, Canada; Samantha Salvi-Cruz, Vanderbilt University Medical Center, Nashville, Tennessee, USA.

## Acknowledgements

Bridge2AI-Voice is funded by the NIH Common Fund through the Bridge2AI program, grant number OT2OD032720. The involvement of JA and BW in this work was also supported by the NIH Center for Interventional Oncology and the Intramural Research Program of the National Institutes of Health, National Cancer Institute, the National Institute of Biomedical Imaging and Bioengineering, via intramural NIH Grants Z1A CL040015 and 1ZIDBC011242, and the NIH Intramural Targeted Anti-COVID-19 (ITAC) Program, funded by the National Institute of Allergy and Infectious Diseases. The participation of HH was made possible through the NIH Medical Research Scholars Program, a public-private partnership supported jointly by the NIH and contributions to the Foundation for the NIH from the Doris Duke Charitable Foundation, Genentech, the American Association for Dental Research, the Colgate-Palmolive Company, and other private donors. DAC was supported by the Pandemic Sciences Institute at the University of Oxford; the National Institute for Health Research (NIHR) Oxford Biomedical Research Centre (BRC); an NIHR Research Professorship; a Royal Academy of Engineering Research Chair; the Wellcome Trust funded VITAL project (grant 204904/Z/16/Z); the EPSRC (grant EP/W031744/1); and the InnoHK Hong Kong Centre for Cerebro-cardiovascular Engineering (COCHE).

## Disclosures / Conflicts of Interest

The content of this manuscript does not necessarily reflect the views, policies, or opinions of the National Institutes of Health (NIH), the U.S. Government, nor the U.S. Department of Health and Human Services. The mention of commercial products, their source, or their use in connection with material reported herein is not to be construed as an actual or implied endorsement by the U.S. government nor the NIH.

## Data Availability Statement

Data can be made available upon reasonable requests with raw voice data needing execution of a DUA with the University of South Florida.

## Notes

### Competing Interest Statement

The authors have declared no competing interest.

### Author Declarations

The ethics committee/IRB of the University of South Florida gave ethical approval for this work

